# Aberrant visual salience in participants with schizophrenia during free-viewing of natural images

**DOI:** 10.1101/2022.11.21.22282553

**Authors:** Masatoshi Yoshida, Kenichiro Miura, Michiko Fujimoto, Hidenaga Yamamori, Yuka Yasuda, Masao Iwase, Ryota Hashimoto

**Author notes:** Correspondence (M.Y.), (K.M.).

## Abstract

Abnormalities in visual exploration affect the daily lives of patients with schizophrenia; however, its origin is unknown. In this study, we examined whether such abnormalities reflect aberrant processing of visual salience. Eye movements of 82 patients and 252 healthy individuals viewing natural and/or complex images were examined using saliency maps for static images to determine the contributions of low-level visual features to salience-guided eye movements. The results showed that the gazes of the participants with schizophrenia were attracted to position in the images with high orientation salience but not luminance or color salience. Further analyses revealed that orientation salience defined by the L+M channel of the DKL color space is specifically affected in schizophrenia, suggesting abnormalities in the magnocellular visual pathway. These results suggest aberrant processing of visual salience in schizophrenia, thereby connecting the dots between abnormalities in early visual processing and the aberrant salience hypothesis of psychosis.

---

Schizophrenia is a mental disorder with psychosis, that is, delusion and hallucination. Previous studies have successfully identified various behavioral markers of schizophrenia ^1^. However, to understand the pathological mechanisms of schizophrenia, it is necessary to find links between these behavioral markers and theories about how the symptoms develop. Here, we examined the relevance of exploratory eye movements as a behavioral marker and the aberrant salience hypothesis of psychosis ^2–4^, which states that aberrant assignment of salience to elements of one’s experience leads to delusion and hallucination.

Various abnormalities in eye movements have been reported in patients with schizophrenia ^5,6^. Among them, significant differences in eye movements are observed in a simple free-viewing task. Gaze covers a relatively large area of the images in healthy participants, whereas participants with schizophrenia tend to limit their gaze to a narrower area of the images, resulting in shorter scanpath lengths than healthy participants ^7–10^. Visual and visuo-cognitive processing are affected ^11–13^, while the motor aspects of saccadic eye movements are less affected ^10,14^. In schizophrenia, it would be reasonable to assume that abnormalities in visual exploration result from abnormalities in visual or visuo-cognitive processing.

We sought to determine whether these abnormalities in eye movements were due to aberrant assignment of visual salience to objects in the visual scene. To this end, we examined eye movements during free-viewing of natural images in participants with schizophrenia (SZ) and healthy controls (HC). The visual salience of the presented images was quantified by Itti and Koch’s computational model of salience, which has already proven useful for predicting and analyzing the visual exploration behavior of humans and non-human primates ^15–20^. In the present study, we first tested whether the gaze sequences of participants with schizophrenia differed from those of healthy controls in terms of salience values. Since the results showed case-control differences, we examined in detail which low-level visual features contributed to the differences in eye movements and found that the difference stemmed specifically from orientation salience. We then explored the origin of the effects of orientation salience by examining the stages of the salience computation in the model.

## Results

### Visual-oculomotor properties are affected in schizophrenia but oculomotor properties are not

Eye-tracking data was obtained from 82 participants with schizophrenia and 252 healthy controls while freely viewing 56 natural and/or complex images. Supplementary Table 1 summarizes the demographics of the participants, scores of cognitive tests, and saccade characteristics during free-viewing. Analysis of saccades revealed that visual exploration is affected in schizophrenia, as reported in previous studies ^10,14^. The average number of saccades during 8 seconds of the viewing period, the average amplitude of saccades, and the scanpath length were smaller in participants with schizophrenia than in healthy controls (Supplementary Table 1). In contrast, oculomotor properties, as assessed by the fitted parameters of the main sequence relationship, were relatively unaffected (Supplementary Table 1).

### Salience-guided eye movements are affected in examples of single images

To investigate the relationship between the gaze positions (defined as endpoints of regular saccades) and the visual salience of the test images, saliency maps were computed using the Itti-Koch saliency computational model (Fig. 1A) ^15,21^.

**Fig. 1.**
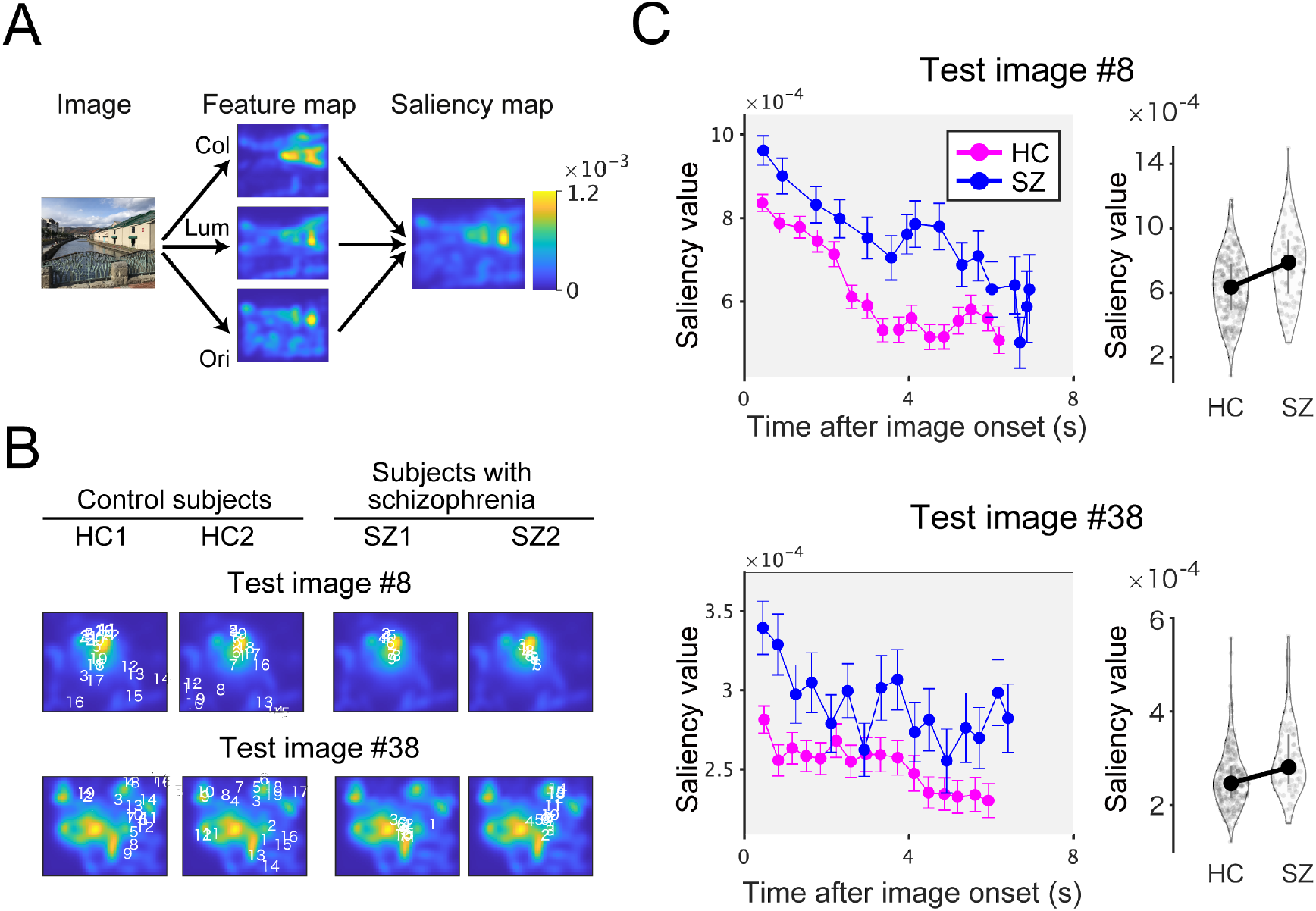
Salience-guided eye movements are affected in examples of single images. (A) The saliency map was calculated from the Itti-Koch model. Visual salience for low-level visual features (color, “Col”; luminance, “Lum”; orientation, “Ori”) was also computed in this model. (B) Gaze positions of two control subjects (left) and two SZs (right) represented by numbers and superimposed on the saliency map of test images. Numbers indicate saccade order. (C) Left: The saliency values averaged across participants are plotted over time for test images #8 (top) and #38 (bottom). Error bars denote standard errors across participants. Magenta, HC; blue, SZ. Right: the saliency values averaged across participants and saccades plotted as a violin plot. Symbols denote the median. Error bars denote the first and the third quartile.

In representative examples (for the test image #8 and #38), saccades of healthy controls were distributed not only in highly salient positions (shown in yellow) but also in other locations in the image (Fig. 1B, left). In contrast, saccades of participants with schizophrenia remained at salient locations of the image during free-viewing (Fig. 1B, right). To quantify the time course of salience values during the 8-second viewing period, the salience values for each gaze were averaged across participants for the test images and were plotted across time (Fig. 1C, left). The mean salience values were higher throughout the viewing period in participants with schizophrenia (SZ; blue in Fig. 1C, left) than in healthy controls (HC; magenta in Fig. 1C, left). The difference between the participant groups becomes more evident when the salience values were averaged across all saccades during the viewing period (Fig. 1C, right).

### Visual salience attracts the gaze of participants with schizophrenia

To assess population averages, we plotted the salience values for each saccade averaged across all test images and all participants (Fig. 2A). Consistent with the single image results (Fig. 1C), the mean saliency values were consistently higher in SZs than in HCs.

**Fig. 2.**
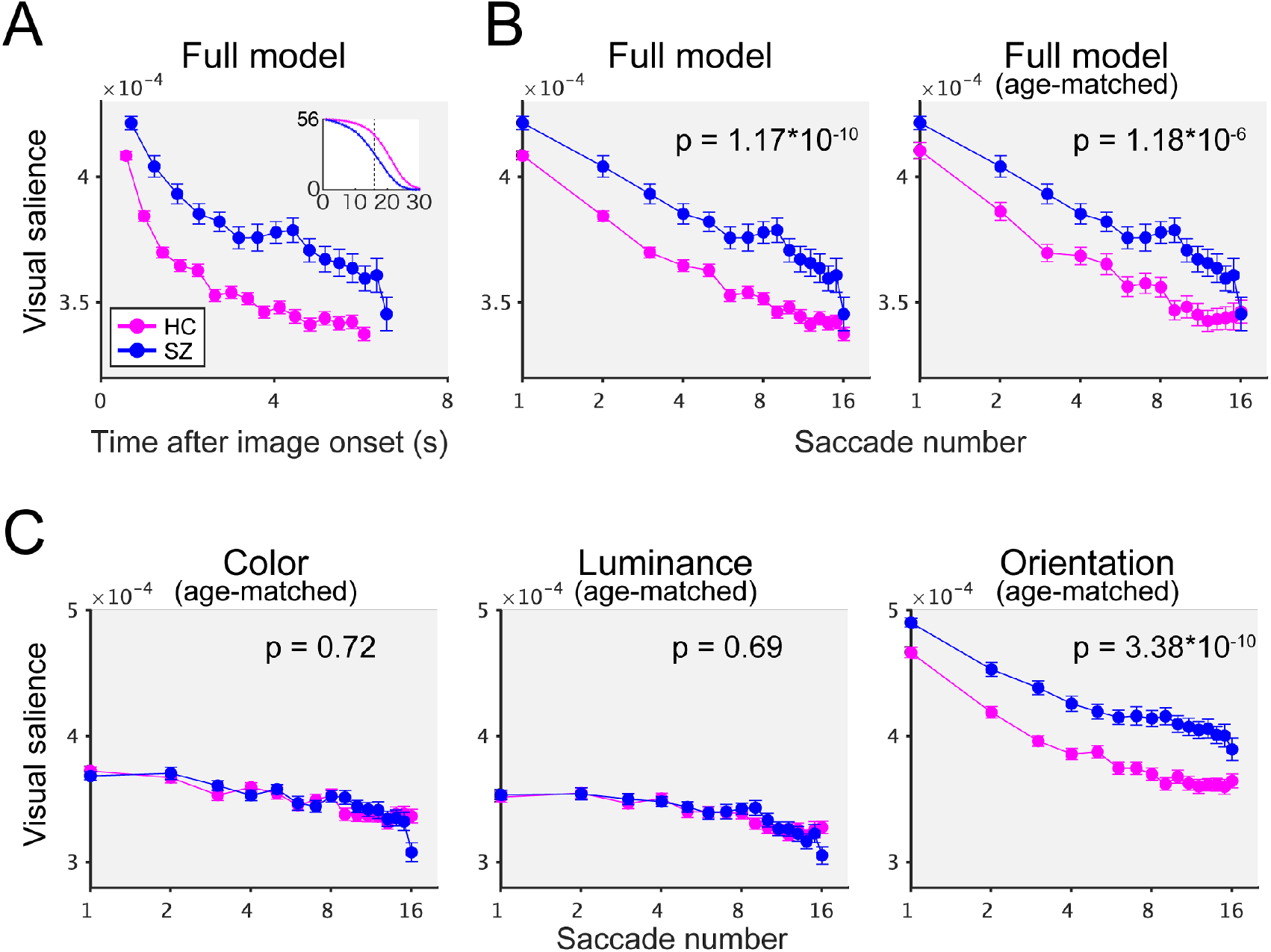
The gaze of participants with schizophrenia is attracted by orientation salience. (A) The saliency values averaged across test images and participants are plotted over time after image onset. Magenta, the healthy controls (HC; n=252); blue, the participants with schizophrenia (SZ; n = 82). Each point denotes the mean salience value for each saccade (1st through 16th). Error bars denote the standard errors across participants. Inset: number of images obtained for each number of saccades (up to 56) during the 8-sec viewing period. The dotted line denotes the cutoff point (16th saccades). See Methods for details. (B) (Left) Same as (A) but the data are plotted across saccade numbers on a log scale. (Right) Age-matched resampled data are plotted for healthy controls. Numbers on the plots denote P values for the main effect of the participant group. (C) As in (B), but for single feature salience models.

For statistical analyses, a linear mixed model with random intercepts and slopes was used to test the main effect of the participant group (SZ vs. HC) and the interaction between the participant group and the fixation number. Using the log of saccade numbers as a factor, it is more reasonable to fit the data with a linear mixed model (Fig. 2B, left). Since the interaction term was not significant (F(1, 330.11) = 0.17 and P = 0.67), a model without interaction was selected. The main effect for the participant group was highly significant (F(1, 332.00) = 44.29 and P = 1.17*10^−10^). These results suggest that the gaze of SZs is more likely to be directed toward visually salient locations than that of HCs.

When the data from age-matched resampled data for HC were compared with the data from SZ (Fig. 2B, right), the results of the statistical analysis were not affected. The interaction term was not significant (F(1, 159.69) = 0.02 and P = 0.87), and a model without interaction was selected. The main effect for the participant group was highly significant (F(1, 162.00) = 25.49 and P = 1.18*10^−6^). The resampling procedure was repeated 100 times, one of which was used throughout the following analyses. See also the supplementary text, “Consideration of resampling scheme” for more details.

### The gaze of participants with schizophrenia is attracted by orientation salience

To assess the contribution of low-level visual features, single-feature saliency maps for color (“Col”), luminance (“Lum”), and orientation (“Ori”) were used for analysis as in the full salience model (Fig. 1A). In all three models, interaction terms were not statistically significant (F(1, 160.79) = 0.93 and P = 0.34 for color, F(1, 161.60) = 0.52 and P = 0.47 for luminance, and F(1, 160.14) = 1.79 and P = 0.18 for orientation, respectively). In the color (Fig. 2C left) and the luminance (Fig. 2C middle) models, the main effects for the participant group were not significant (F(1, 162.01) = 0.16 and P = 0.69 for color and F(1, 161.97) = 0.12 and P = 0.72 for luminance, respectively). In contrast, the main effect of the participant group was highly significant (F(1, 161.99) = 44.80 and P = 3.39 *10^−10^) in the orientation model (Fig. 2C right). These results suggest that the main effect in the participant group in the full model (Fig. 2B) is explained sorely by orientation salience.

### The specific effect of orientation salience is also evident in time-averaged data

Since the liner mixed model showed no significant interaction effect between the participant group and saccade numbers in all four salience models (Fig. 2B and C), it is reasonable to summarize the data by averaging all saccades during the viewing period (Fig. 3A). The mean saliency value from the full model was higher in SZ than in HC (Z = 4.82, P = 5.75 *10^−6^; in Wilcoxon rank-sum test with Bonferroni correction). The effect size evaluated by Cliff’s delta was 0.44. The mean salience value from the orientation model was higher in SZ than in HC (Z = 6.41, P = 5.64 *10^−10^; in Wilcoxon rank-sum test with Bonferroni correction). The effect size evaluated by Cliff’s delta was 0.58, which is classified as a large effect. On the other hand, the mean salience value from the color and the luminance models were not significantly different between SZ and HC (Z = 0.08, P > 1.0 for color; Z = 0.26, P > 1.0 for luminance; in Wilcoxon rank-sum test with Bonferroni correction). Based on these findings, the following analyses were performed on the time-averaged data.

**Fig. 3.**
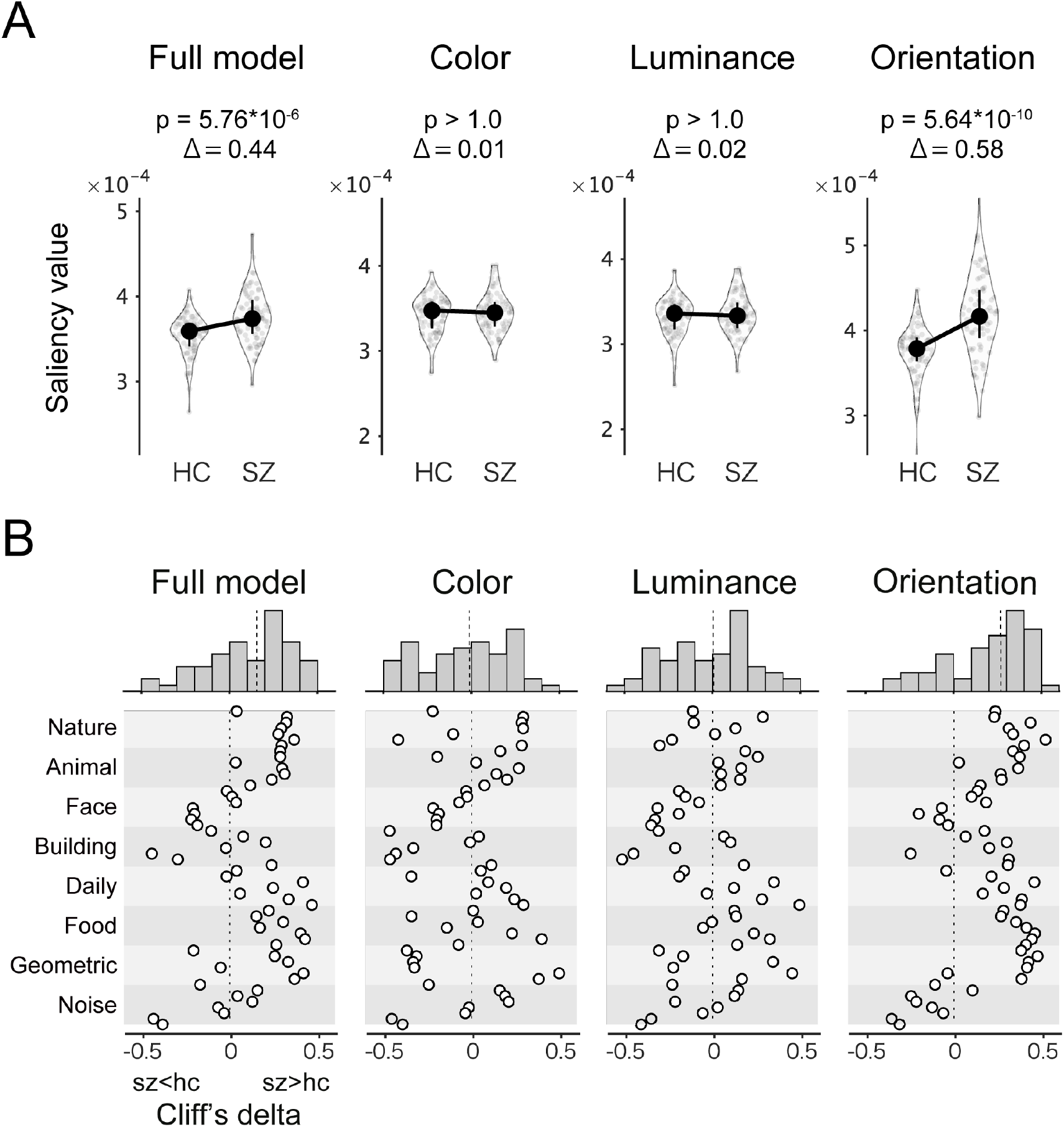
Image category does not affect the effect of orientation salience. (A) Mean saliency values for all images and saccades are plotted for the healthy controls (HC) and the schizophrenia group (SZ). Data from four salience models are plotted. Symbols denote median values. Error bars denote the first and the third quartile. Numbers on the plots denote P values (“p”) and the effect sizes (“Δ”, Cliff’s delta) of the Wilcoxon rank-sum test. (B) Effect sizes (Cliff’s delta) for the test of difference in time-averaged saliency values. Top: histograms of effect sizes.

All of these mean salience values were significantly higher than chance. See also the supplementary text, “Consideration of random sampling scheme” for more details. We also tested whether the saliency values for the first saccade were significantly different between SZ and HC (Supplementary Fig. 1). See Discussion for details.

### Image category does not affect the effect of orientation salience

To examine whether the effects of orientation salience depend on image category, the effect size (Cliff’s delta) that evaluates the difference between the time-averaged saliency values of HC and SZ for each test image was plotted for all four salience models (Fig. 3B). Positive values in the effect size indicate that saliency values were higher in SZ than in HC. The results (Fig. 3B) show that the effect sizes for the Full and the orientation models were consistently positive for all image categories except for the Face and Noise categories. This suggests that the effect of orientation salience is overall robust to image categories.

### The key computational stage that produces differences in orientation salience is Gabor filtering

Next, we examined which stage of the salience computation produces differences in orientation salience. In the Itti-Koch salience model, saliency maps for the orientation feature are generated through five processing stages (Fig. 4A): 1) transforming the input image into a grayscale image with various spatial resolutions (i.e., Gaussian pyramids), 2) filtering the images with Gabor patches with four different orientations, 3) center-surround inhibition, 4) peak normalization, and finally, 5) adding all images together to obtain the saliency map of the orientation feature. These intermediate maps were calculated and used for analysis as was done in the analysis of Fig. 3A. Differences between participant groups were detected not only in the saliency map for the orientation feature (Fig. 4B, rightmost) but also in peak normalization, center-surround inhibition, and Gabor filter (Fig. 4B, middle). In contrast, no differences between participant groups were detected in intensity (Fig. 4B, leftmost). These differences became more evident when each comparison was evaluated by the effect size (Fig. 4C). These results suggest that the differences between participant groups emerge at the stage of Gabor filtering with four orientations, a unique computational stage that is not present in the color or luminance salience but only in the orientation salience.

**Fig. 4.**
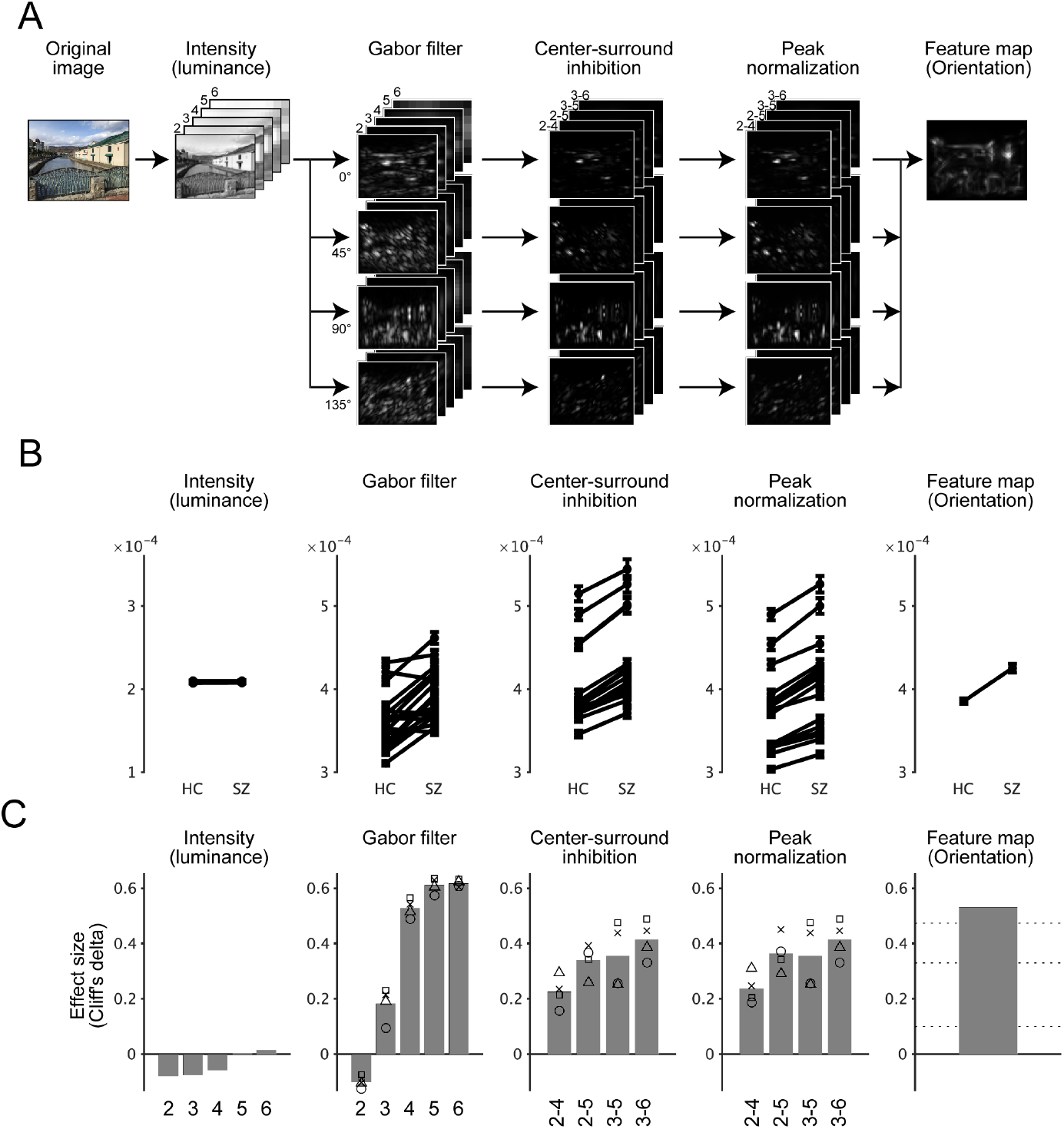
The key computational stage that produces differences in orientation salience is Gabor filtering. (A) The Itti-Koch saliency model computes the saliency map of the orientation feature through five stages. Intermediate images for a given image are shown in grayscale. (B) Averaged map values for intermediate images are plotted as in Fig. 3A. (C) Effect sizes (Cliff’s delta) for the tests in (B). Numbers indicate the spatial scales of the Gaussian pyramids. In the three plots in the center, the mean effect size for four orientations (bar), as well as the effect size for four orientations (symbols) were plotted. “o,” 0 degrees; “x,” 45 degrees; triangle, 90 degrees, and square, 135 degrees. The horizontal lines in the rightmost plot (“Saliency map (Orientation)”) indicate the common guidelines for the effect size (Cliff’s delta). See Methods for details.

It is also interesting to note that the effect size of Gabor filtering is larger for low-pass images (those with higher numbers in Fig. 4C). Since it is already known that the magnocellular pathway conveys relatively low spatial frequency signals ^22^ and that the magnocellular pathway is specifically impaired in schizophrenia ^11^, this point is directly examined in the next section.

### the L+M channel of the DKL color space is dominant in the effect of orientation salience

In the Itti-Koch salience model (hereafter referred to as the “original model”), the orientation salience is calculated from grayscale images. Instead, it is possible to decompose the test images into images of three channels (L+M, L-M, and S-(L+M)), based on the DKL color space ^23^. The decomposed images were then processed to obtain saliency maps with (orientation map) or without (intensity map) Gabor filtering (Fig. 5A). We refer to this as the “Extended six-channel model.”

**Fig. 5.**
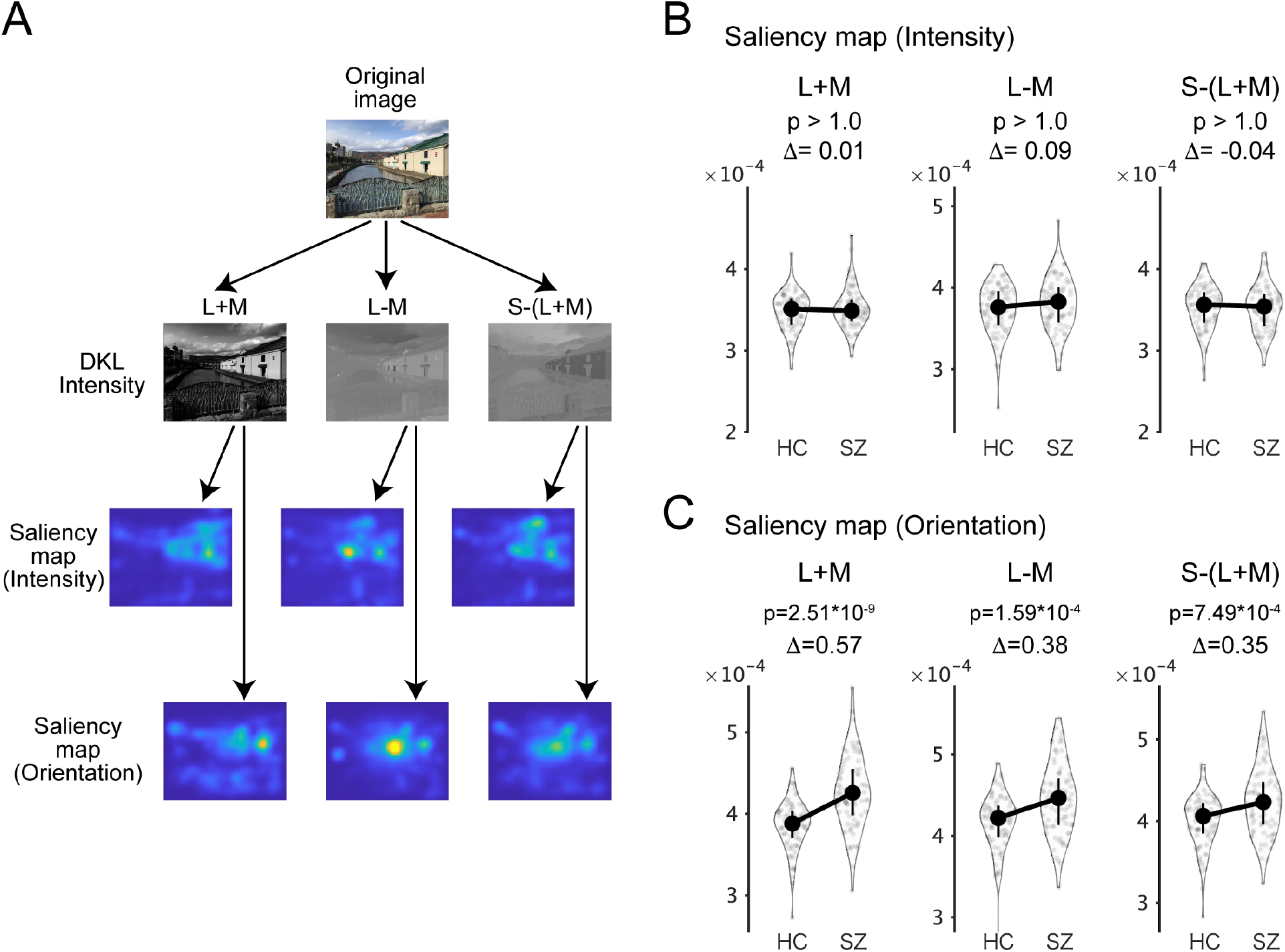
the L+M channel of the DKL color space is dominant in the effect of orientation salience. (A) To construct the extended six-channel model, the original image was decomposed into three channels in the DKL color space: the magnocellular L+M channel, the parvocellular L-M channel, and the koniocellular S-(L+M) channel. Then saliency maps for intensity and orientation were obtained for each of the three channels (six maps in total). (B and C) As in Fig. 3A, the salience values averaged across test images and saccades were plotted, but for six saliency maps. Symbols are the same as in Fig. 3A.

Then the salience values for these six maps were compared between SZ and HC (Fig. 5B and C). In the intensity maps (Fig. 5B), all differences were not statistically significant (Z = 0.21, P > 1.0 for the L+M channel; Z = 1.09, P > 1.0 for the L-M channel; Z = -0.51, P > 1.0 for the S-(L+M) channel; in Wilcoxon rank-sum test with Bonferroni correction). In the orientation maps (Fig. 5C), all of the differences were statistically significant (Z = 6.25, P = 2.51 *10^−9^ for the L+M channel; Z = 4.20, P = 1.59 *10^−4^ for the L-M channel; Z = 3.84, P = 7.49 *10^−4^ for the S-(L+M) channel; in Wilcoxon rank-sum test with Bonferroni correction). These results further support the finding of a specific effect of orientation salience and suggest that the L+M channel of the DKL color space is dominant in the effect of orientation salience.

### Orientation salience is correlated with the scores of cognitive tests and visual oculomotor characteristics

Next, we examined whether orientation salience is correlated with demographic data, cognitive ability, and eye movement characteristics collected from the same participants (See ^10,24^ for details). When the dependent variables (demographic data, cognitive tests, or eye movement characteristics) were fitted with orientation salience with the participant group as a covariate, age was not correlated with orientation salience but WAIS-3, processing speed (PS), social functioning scale (SFS), and scanpath length (SPL) were correlated with orientation salience (Fig. 6A).

**Fig. 6.**
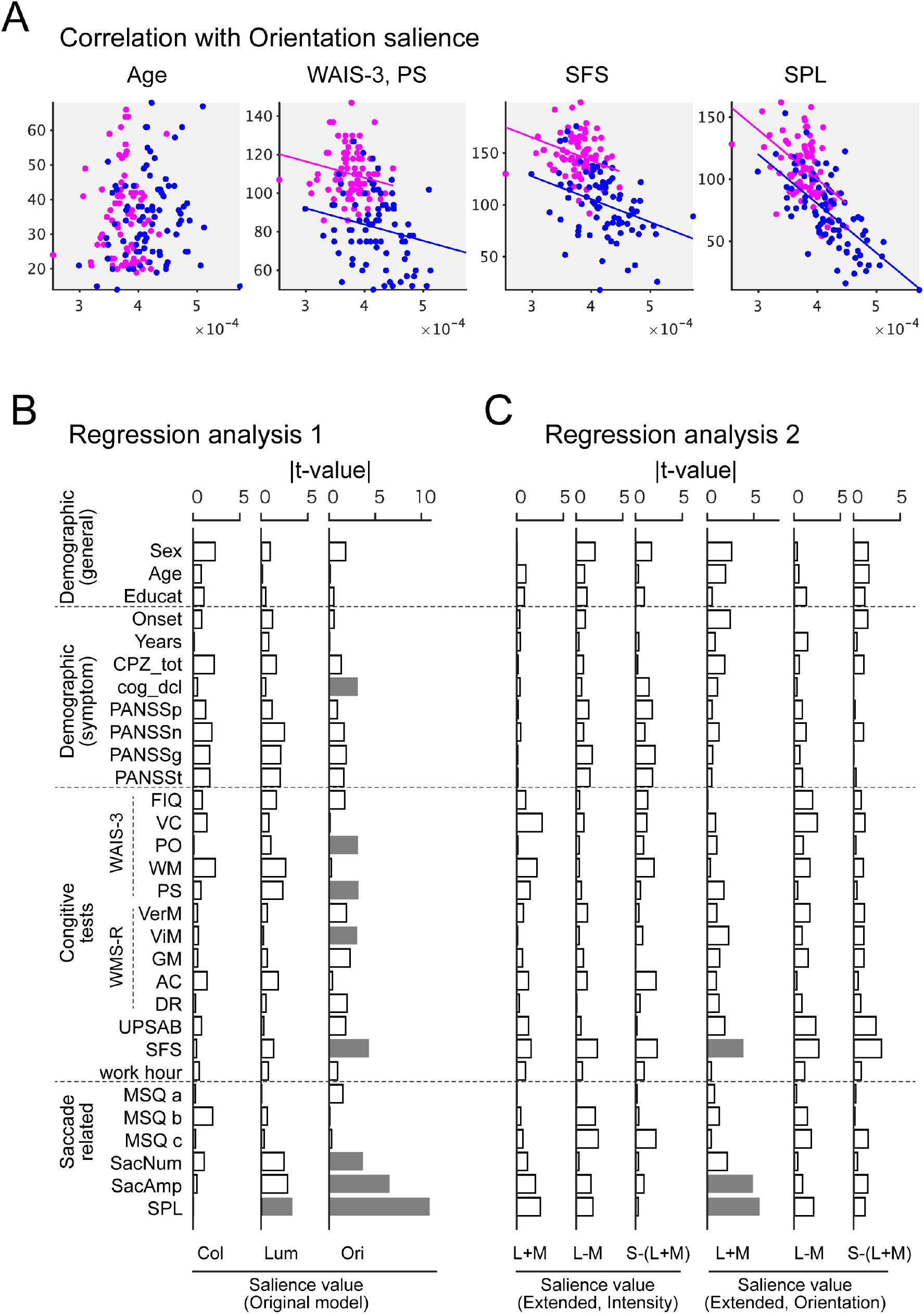
Orientation salience is correlated with the scores of cognitive tests and visual oculomotor characteristics. (A) Scattered plots for the mean orientation salience (averaged across images and saccades) and age, WAIS-3 processing speed (PS), social functioning scale (SFS), and scanpath length (SPL). Each dot represents the value for one participant. Magenta: healthy controls (HC); blue, participants with schizophrenia (SZ). Lines indicate regression lines for each participant group. (B) Absolute value of t-values from regression analysis 1, where the dependent variables were fitted individually with the saliency values of the original model. Gray bars indicate statistical significance (P < 0.05) after correction of multiple comparisons by FDR. (C) As in (B), but those from regression analysis 2, where the dependent variables were fitted individually with the saliency values of the extended six-channel model.

For a more systematic analysis, two regression models were constructed. In the first model, the dependent variables were fitted with saliency values from the original model, with the participant group as a covariate. The t-values obtained from the fitting were then plotted (Fig. 6B). Orientation salience is correlated with various scores of cognitive tests and eye movement characteristics (indicated by the gray bars in Fig. 6B). Regression analysis also suggested that these correlations are specific to orientation.

In the second model in which the dependent variables were fitted with saliency values from the extended six-channel model, with the participant group as a covariate, orientation salience in the L+M channel was significantly correlated with a cognitive test score (SFS) and eye movement characteristics (saccade amplitude and SPL) (Fig. 6C). In contrast, orientation saliences in the other channels are not correlated with these values. These results again suggest that the L+M channel is dominant in the effect of orientation salience.

### Saliency maps for orientation are correlated with fixation maps

In Fig. 3A and Fig. 5C, saliency values were evaluated separately for each feature/channel. It is important to note that saliency maps for the different features/channels are spatially correlated (Fig. 7B). This correlation may affect statistical analyses in Fig. 3A and Fig. 5C. To address this issue, we calculated the partial correlation between the saliency maps and the “Fixation maps” which quantifies gaze distribution as a density map summed across participants for each test image (Fig. 7A). We also created a fixation map for the difference between SZ and HC (“SZ-HC”) (Fig. 7A). The partial correlations of these maps were then calculated for each test image (n=56).

**Fig. 7.**
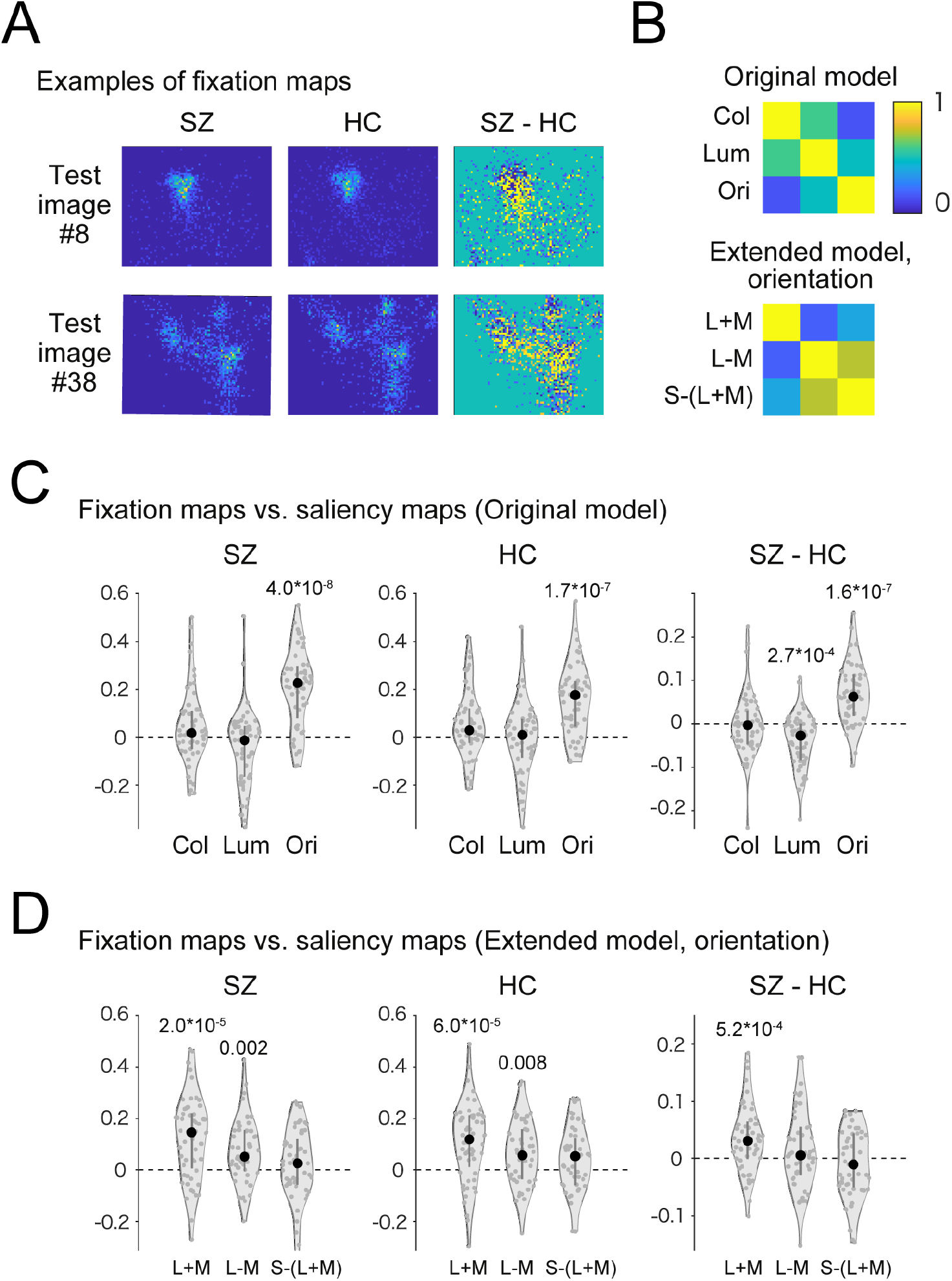
Saliency maps for orientation are correlated with fixation maps. (A) Examples of fixation maps for test images #8 and #38. SZ, the participants with schizophrenia; HC, the healthy controls (age-matched resamples); SZ-HC, the difference between SZ and HC. (B) (Top) Colors indicate the median of partial correlation coefficients between saliency maps of the original model (top) and the extended six-channel model (bottom). (C) Violin plots of the partial correlation between the fixation maps and saliency maps (original model). Symbols are the same as in Fig. 3A. (D) Violin plots of the partial correlation between the fixation maps and saliency maps (the extended six-channel). Only the correlations with orientation maps are shown. Numbers in (C) and (D) indicate P values for a significant difference from zero (Wilcoxon signed-rank test after Bonferroni correction). Only the P values less than 0.05 are shown.

For the original model, we plotted the partial correlations between the fixation maps and the saliency maps for the three features (color, luminance, and orientation) (Fig. 7C). For the three fixation maps (SZ, HC, and SZ-HC), the partial correlation with the saliency map for orientation was significantly larger than zero (P = 4.0*10^−8^ for SZ, P = 1.7*10^−7^ for HC, and P = 1.6*10^−7^ for SZ-HC, respectively; Wilcoxon signed-rank test with Bonferroni correction). In contrast, the partial correlations with the saliency map for color and luminance were not significantly larger than zero (P > 0.05, Wilcoxon signed-rank test with Bonferroni correction) except for a negative correlation in luminance for SZ-HC (P = 2.7*10^−4^).

For the extended six-channel model, three maps for orientation were shown (Fig. 7D). For two fixation maps (SZ, HC), the partial correlations with the saliency map for the L+M and the L-M channels were significantly larger than zero (P = 2.0*10^−5^ and 0.002 for SZ, P = 6.0*10^−5^ and 0.008 for HC; Wilcoxon signed-rank test with Bonferroni correction). On the other hand, for the SZ-HC fixation map, the partial correlation with the saliency map of the L+M channel was significantly larger than zero (p = 5.2*10^−4^, Wilcoxon signed-rank test with Bonferroni correction). These results suggest that the L+M channel contributes specifically to the effect of orientation salience, even after accounting for spatial correlations between saliency maps.

## Discussion

In this study, we examined whether abnormalities in eye movements result from aberrant processing of visual salience. For this purpose, we analyzed the eye movement data during free-viewing with Itti-Koch’s salience model (Fig. 1). We found that the saliency values at the gaze of SZs were persistently higher during the viewing period compared to the HCs (Fig. 2A and B). Further analysis using single-feature saliency maps revealed that this difference was due to orientation salience (Fig. 2C). We then confirmed that these results are robust for various image categories (Fig. 3). Next, we delved into the stages of salience computation and found that differences between SZs and HCs were found in an early stage of salience computation, where grayscale images were filtered with Gabor patches with four orientations (Fig. 4). We also analyzed the gaze with an extended salience model that evaluates the channels for the DKL color space separately and found that the L+M cannel has a dominant role in the effects of orientation salience (Fig. 5). In addition, the saliency values at gazes were not correlated with symptom-related measures such as PANSS scores but were correlated with various measures of cognitive functions and saccade-related characteristics (Fig. 6). Finally, we evaluated a spatial correlation between fixation maps and saliency maps and found that orientation salience in the L+M channel correlates specifically with the difference between the fixation maps of the SZs and those of HCs. These results suggest that visual processing of orientation salience is affected in participants with schizophrenia, which is consistent with the aberrant salience hypothesis of psychosis, as will be discussed shortly.

### Relevance to the aberrant salience hypothesis of psychosis

The aberrant salience hypothesis of psychosis proposes that an aberrant assignment of salience to the elements of one’s experience leads to delusion and hallucination ^2–4^. The concept of salience in this hypothesis covers not only salience derived from emotion and motivation (motivational/incentive salience) ^25^, but also salience due to novelty and sensory features (perceptual salience) ^26^. The present study provides direct evidence that visual salience is affected in schizophrenia, thereby explicitly extending the concept of aberrant salience to the visual domain. In support of our findings, a recent study revealed that brain responses to images with various forms of salience such as novelty, negative emotion, targetness, and rarity/deviance are affected in schizophrenia ^27^.

### Relationship with eye movement abnormalities: visual-oculomotor properties

As described in the Introduction, visual-oculomotor properties such as scanpath length, saccade number, and saccade amplitudes are affected in the gaze of SZs during free-viewing (see also Supplementary Table 1). We found that these visual-oculomotor properties are correlated with saliency values (Fig. 6). Since the bottom-up salience of the images is related to target selection, the abnormalities in visual exploration that have been reported in various studies ^11^ arise, at least in part, from the affected salience-guided eye movement found in this study.

### Relationship with eye movement abnormalities: inhibition-of-return

Recently, our research group reported the finding that inhibition-of-return is impaired in SZs ^28^. This might raise the question of whether return saccades toward salient stimuli are more frequent in schizophrenia (Supplementary Fig. 1A). If this is the case, salience values at the gaze could be higher in SZ than in HC. However, it is unlikely because salience values at the first gaze for the viewing period, where it has no contribution to inhibition-of-return, were significantly higher in SZ than HC in the orientation model (Supplementary Fig. 1C). Furthermore, if the SZs made more return saccades toward salient positions than in the HCs, the slope of the time course of saliency values would be shallower in the SZs than in the HCs (Supplementary Fig. 1B, left). However, this is not the case because analyses using linear mixed models showed no interaction between the participant group and saccade numbers. Thus, abnormalities in visual salience found in this study have a different origin from abnormal inhibition-of-return. For further analysis and discussion, see also Okada et al., 2021^28^.

### Contribution of low-level visual features, early visual cortex, and the magnocellular pathway

We found that salience computation for the orientation feature is specifically affected in schizophrenia. Consistent with this finding, previous findings have also shown that orientation processing is affected in schizophrenia ^29,30^. Other papers have shown that the early visual cortex is involved in changes in contextual modulation in schizophrenia ^31–33^. The effect of orientation salience can explain impairment in contour integration in schizophrenia ^30^ because contour integration can be distracted by aberrant processing of orientation salience.

Based on the analysis of Fig. 4 and the accompanying text, we argue that the crucial difference between SZs and HCs stems from their response to Gabor-filtered images, which resembles responses in the early visual cortex. On the other hand, there was no difference in gazes toward luminance salience. Computation of luminance salience involves center-surround inhibition without Gabor filtering. Such processing resembles the response of the lateral geniculate nucleus (LGN). We propose that aberrant orientation salience reflects abnormalities at the level of the primary visual cortex but not at the level of the LGN. Post-mortem morphological studies of the schizophrenia brains support this possibility. The volume and number of neurons in the primary visual cortex are reduced in schizophrenia ^34,35^ but it is not the case for the LGN ^36,37^. Thus, the idea that visual abnormalities in schizophrenia occur between the LGN and primary visual cortex is consistent with both previous studies and the present current study.

The analyses in Figs. 5, 6, and 7 suggest that orientation salience in the L+M channel of the DKL color space is specifically affected in schizophrenia. Since the L+M channel carries information in the magnocellular pathway ^23^, the present results are consistent with previous findings that visual information processing in the magnocellular pathway is specifically affected in schizophrenia ^38–40^.

### Relationship with clinical/cognitive tests

Fig. 6 shows that symptom-related measures such as the PANSS total score, CPZ equivalent, and duration of illness were not significantly correlated with orientation salience. These results suggest that orientation salience is a trait marker rather than a state marker. This is consistent with the view that eye movement-related measures are trait markers of schizophrenia ^41^.

Fig. 6 also shows that scores on some of the cognitive tests are correlated with orientation salience. This is consistent with previous findings that scores of cognitive tests are correlated with visual oculomotor characteristics such as scanpath length^24^. It is unlikely that the abnormality in scanpath length causes abnormalities in orientation salience, as oculomotor properties of eye movements are less affected in schizophrenia. Rather, we propose that aberrant processing of orientation salience causes abnormalities in visual exploration, which can be assessed by visual oculomotor properties such as scanpath length, which in turn causes difficulty in cognitive abilities such as social functioning.

### Visual salience in schizophrenia and autism spectrum disorder (ASD)

Analysis of visual salience during free-viewing can be a useful method for detecting psychiatric or neurological disorders ^42,43^. The current findings added another example of the successful application of the computational salience model to the analysis of eye movements in participants with psychiatric disorders.

Gazes of participants with autism spectrum disorder (ASD) showed a general increase in visual salience, defined by the low-level visual features ^44^. This appears similar to the findings in the current study about schizophrenia but differs in that the present study reported specific effects of the orientation feature. Also interesting is the finding that the effect size of orientation salience was lower in the Face category than in the other image categories in the present study (Fig. 3B). This suggests that faces are relatively ineffective in discriminating between SZs and the HCs, in contrast to the abnormalities in gaze and processing of faces in ASD ^45^.

In terms of the predictive coding/Bayesian brain hypothesis, both schizophrenia and ASD are explained by difficulties in evaluating prediction errors in sensory signals ^46,47^. How to distinguish the two in terms of computational perspective is an ongoing debate. By extending the concept of visual salience to Bayesian surprise ^48^ and treating visual salience as spatial outliers from the current belief on what we see, saliency analysis could help resolve this issue.

### Future projects

In this study, we obtained clues about what is affected in the brain of schizophrenia, for example, Gabor filtering that is presumably performed in the magnocellular pathway of the early visual cortex. To understand exactly how the brain performs such computation, it is necessary to understand the processes at the neuronal level. To this end, neurophysiological studies using animal models of schizophrenia are needed. Since eye-tracking during free-viewing and analysis of visual salience is an experimental paradigm that has been successfully used in non-human primates such as macaque monkeys ^16^ and marmosets ^18,19^, replicating the present results in a non-human primate model of schizophrenia^19,49^ would open the door to understanding the precise brain mechanism of schizophrenia.

## METHODS

### Participants

Eye movement data were sampled from 82 SZs (male, 42; female, 40) and 252 HCs (male, 144; female, 108) as part of a large-scale cohort recruited at Osaka University (Supplementary Table 1) ^10,14,24^. There was an overlap in data with a previous study on eye movement abnormalities in schizophrenia ^10^. All participants were biologically unrelated, were of Japanese descent, and had no history of the ophthalmologic disease, or neurological/medical conditions that could influence the central nervous system. Specific exclusion criteria included atypical headaches, head trauma with loss of consciousness, chronic lung disease, kidney disease, chronic hepatic disease, thyroid disease, active cancer, cerebrovascular disease, epilepsy, seizures, substance-related disorders, or mental retardation ^10,14^.

SZs were recruited from Osaka University Hospital and had been diagnosed by two or more trained psychiatrists according to criteria from the DSM-IV based on the Structured Clinical Interview for DSM-IV (SCID). Estimated cognitive decline was calculated by the methods described by Fujino ^55^. The current symptoms of the SZs were assessed using the Positive and Negative Syndrome Scale (PANSS) ^56^, and daily antipsychotic use was calculated using chlorpromazine (CPZ) equivalents (mg/day) ^57^.

HCs were assessed for psychiatric, medical, and neurological concerns using a non-patient version of the SCID to exclude individuals with current or past contact with psychiatric services or who had received psychiatric medication.

All participants provided written consent to the study after a full explanation of the study procedures. Anonymity was preserved for all participants. The study was performed in accordance with the World Medical Association’s Declaration of Helsinki and was approved by the Research Ethical Committee of Osaka University, the National Center of Neurology and Psychiatry, and Center for Experimental Research in Social Sciences, Hokkaido University.

### Task and stimuli

In the free-viewing task, the participants faced a 19-inch liquid crystal display monitor (1280 × 1024 pixels) placed 70 cm from the observer’s eyes. Presentation of the visual stimuli was done using the Psychophysics Toolbox extension ^52^ in MATLAB (The Mathworks, Natick, MA, USA). Each trial began with the presentation of a fixation point on the center of the display. Once the participant fixated on the fixation spot for a random time, a test image was presented for 8 seconds. The participant was instructed to view the image as they like. One task consisted of 56 images, the order of which was randomly shuffled for each participant. The images were chosen from eight categories: natural environments, buildings, everyday items, foods, faces, animals, fractal patterns, and noise (seven images for each). The images of natural environments and animals were selected from the International Affective Pictures System (IAPS) ^50^, and the face images from Matsumoto and Ekman ^51^. Since IAPS images are not allowed to be published in scientific journals, only the saliency maps for these images (#8 and #38) are shown in the figures. In cases when images needed to be shown as examples (Figs. 1A, 5A, and 6A), a photograph taken by one of the authors was used.

### Recording and preprocessing of eye movement data

Recording and preprocessing of eye movement data were performed as follows ^10^. Eye position and pupil area of the left eye were measured at 1 kHz using EyeLink1000 (SR Research, Ontario, Canada). Eye position data (in degrees) were smoothed with a digital FIR filter (−3 dB at 30 Hz), and eye velocity and acceleration traces were derived from a two-point difference algorithm. Eye movement recordings were segmented into blink, saccade, and fixation periods. Detected saccades included both regular saccades and microsaccades. Here, following previous papers on microsaccades during free-viewing such as ^58^, saccades with amplitudes greater than one degree were selected as regular saccades.

### Computational models and saliency analysis

To assess salience-guided eye movements, we used a validated computational model of visual attention and compared it with individual eye movements. The saliency maps for the test images were computed with the Itti-Koch saliency model for static images ^15^ implemented in the GBVS toolbox for Matlab ^21^. The Itti and Koch model is a neurobiologically inspired model which computes salient locations for low-level visual features (Fig. 1A). Since we are interested in neurobiological mechanisms of saliency-guided eye movements, the Itti-Koch model was chosen.

In the full model, saliency maps of the three features were summed with equal weights. When evaluating the contribution of low-level visual features, single-feature saliency maps were used (Fig. 1A). For a test image of 640*512 pixels, we obtained a saliency map of 80*64 pixels. To treat the saliency maps as density maps, all maps were normalized so that the sum of saliency values of all pixels was one.

Intermediate files for salience computation were used for the analysis in Figs. 4 and the accompanying text. These files were also generated by the GBVS toolbox.

### Extended six-channel model for visual salience

An “extended six-channel model” for visual salience was constructed for the analysis in Figs. 5, 6, and 7 and the accompanying text. For this purpose, the test images were decomposed into three-channel images (L+M, L-M, and S-(L+M)) based on the DKL color space ^23^. The decomposed images were then subjected to salience computation separately with (orientation map) or without (intensity map) Gabor filtering (Fig. 5A). The luminance salience in the original model corresponds approximately to the intensity map of the L+M channel. The color salience in the original model corresponds approximately to the intensity map of the L-M channel plus the S-(L+M) channel. The orientation salience in the original model corresponds approximately to the orientation map of the L+M channel. The other two maps (the orientation map of the L-M channel and the orientation map of the S-(L+M) channel) are new components not found in the original model.

### Age-matched resampling

To account for the effect of the age difference between the healthy control group (HC, 28.8 ± 11.5 years, mean ±SD) and the schizophrenia group (SZ, 35.1 ± 12.4 years, mean ±SD), resampling of the data for the HC group was performed to match the mean age of the HC participants with that of the SZ group. For this purpose, the ages of both groups were grouped into 5-year bins. The HC participants in each bin were then randomly selected to match the number of SZ participants in each bin. This procedure was repeated 100 times to obtain 100 sets of resampled HC participants. The mean age of the resampled HC participants ranges from 34.9 – 35.2 years, in close agreement with that of the SZ group (35.1 years). The standard deviation of the age of the resampled HC participants ranges from 12.0 – 12.5 years, in close agreement with that of the SZ group (12.4 years old). We confirmed that the results of statistical analysis were not affected by the choice of the resampled dataset (see supplemental text, “Consideration of resampling scheme”). Therefore, we selected one of the resampled datasets and used it throughout our analysis.

### Statistical analysis

Statistical analyses were performed using MATLAB (Mathworks, NY), except for the linear mixed models, which were performed by the R packages. The significance level was set at p < 0.05. Non-parametric Wilcoxon rank-sum tests after Bonferroni correction were performed in a post-hoc analysis to compare SZ and HC for the median salience values or other indices (Fig. 3A, Fig. 5B and C, Supplementary Fig. 1C). Cliff’s delta (Δ) was used to quantify the effect size of the estimated difference. For interpretation, we followed general guidelines: negligible for |Δ| < 0.147, small for 0.147 <= |Δ| < 0.33, medium for 0.33 <= |Δ| < 0.474, and large for |Δ| >= 0.474^59^.

### Linear mixed models

The procedure for the statistical analysis of the time course of the mean salience values at saccade endpoints (Figs. 1 and 2) is as follows. First, a cut point for the number of saccades was set because the number of saccades during the 8-second viewing time varied between trials and between the participant groups (see Supplementary Table 1 for the mean numbers of saccades; 16.1 in SZ and 20.9 in HC). The cut point was set to 16 because the mean number of images obtained for each saccade number (up to 56) is less than half (Fig. 2A, inset).

We then fit linear mixed models using the R package lme4 ^53^ to obtain estimates and statistics. For this purpose, the salience values for each saccade number and each participant were averaged over 56 test images. Saccades that landed off-screen were treated as not a number. The mean salience values (“Sal”) were then treated as the dependent variable and as a linear function of participant groups (HC and SZ as “SubjectGroup”) and saccade numbers (1st, 2nd…, 16th as “SaccadeNum”). The random effects were modeled using random intercepts and random slopes for individual differences (ranging from 1 to 334 as “SubjectID”) nested under the participant group. The model formulae in R format are as follows:

1. Sal ∼ SubjectGroup + SaccadeNum + (SaccadeNum |SubjectID)
2. Sal ∼ SubjectGroup * SaccadeNum + (SaccadeNum |SubjectID)

Model #1 is a model with the main effects of participant group and saccade number. Model #2 is a model with an interaction between the participant group and saccade number. The two-sided probability values and degrees of freedom associated with each statistic were then determined using the Satterthwaite approximation implemented in the R package LmerTest ^54^.

### General linear models

To examine whether the salience values are corelated to other measures such as demographic data, cognitive tests, and eye movement properties, general linear models were constructed (Fig. 6). The model formulae in R format are as follows:

1. Dependent variables ∼ SubjectGroup + Sal(Col) + Sal(Lum) + Sal(Ori)
2. Dependent variables ∼ SubjectGroup + Sal(Int, L+M) + Sal(Int, L-M) + Sal(Int, S-(L+M)) + Sal(Ori, L+M) + Sal(Ori, L-M) + Sal(Ori, S-(L+M))

In Model #1, the dependent variables (demographic data, cognitive tests, and saccade-related properties) were fitted with the participant group (“SubjectGroup”), and the salience values were calculated by the original saliency model consisting of three features. In Model #2, the dependent variables were fitted with the participant group (“SubjectGroup”) and the salience values were calculated by the extended six-channel model. P values were adjusted for multiple comparisons using FDR.

## Data Availability

All data reported in this study cannot be deposited in a public repository because of ethical prohibitions. To request access, contact the lead contact.

## Code availability

Custom-written codes used for the analysis of this study are available from GitHub (https://github.com/pooneil68/saliency_analysis DOI: 10.5281/zenodo.7312329).

## Acknowledgments

This research was supported in part by AMED under Grant Numbers JP19dm0207069 (MY and KM), JP21dk0307103 (RH), JP21wm0425012 (KM and RH), JP18dm0307002 (RH), and JP21uk1024002 (KM and RH); JSPS KAKENHI grant numbers 22H02936 (MY), JP20H03611 (RH), JP19H05467 (RH), JP20K06920 (KM); and Intramural Research Grant (3-1 and 4-6) for Neurological and Psychiatric Disorders of NCNP (KM and RH).

## Author contributions

Conceptualization, M.Y.; Methodology, M.Y. and K.M.; Software, M.Y.; Investigation, M.F., H.Y., Y.Y. and M.I.; Resources, K.M. and R.H.; Data Curation: M.Y., K.M. M.F., H.Y., Y.Y. and M.I; Writing – Original Draft: M.Y.; Writing – Review & Editing: M.Y. and K.M.; Funding Acquisition, M.Y., K.M., and R.H.; Supervision, K.M. and R.H.

