## Supplementary information for "Aberrant visual salience in participants with schizophrenia during free-viewing of natural images"

### Supplementary Text

**Consideration of resampling scheme.** Considering the possibility that resampled data could affect the results of the statistical analysis, the resampling procedure was repeated 100 times in the statistical analysis with the full model using age-matched control data (Fig. 2B, right). For the interaction term, the range of F values was $5.8*10^{-5}$ - 1.07 and that of P values was 0.30 - 0.99. For the main effect for the participant group in the model without interaction, the range of F values was 19.0 - 36.9 and that of P values was $8.4*10^{-9}$ - $2.2*10^{-5}$. Thus, differences in the choice of resampled data do not affect the result of the statistical analysis. Thus, one of the resampled data was selected and used throughout the analysis in the main text.

**Consideration of random sampling scheme.** In Fig. 3A, we also examined whether the mean salience values were higher than the chance level, here defined as the expected value from random sampling, i.e., $1.95 * 10^{-4}$ (= 1 / (80*64) ). In all cases of the four models and two participant groups, they were significantly higher than chance ($p < 10^{-12}$, Wilcoxon signed-rank test with Bonferroni's correction for multiple comparisons). This confirms that in all models and all participant groups, the participants' gazes were directed to the salient locations in the test images.

**Supplementary Figure**

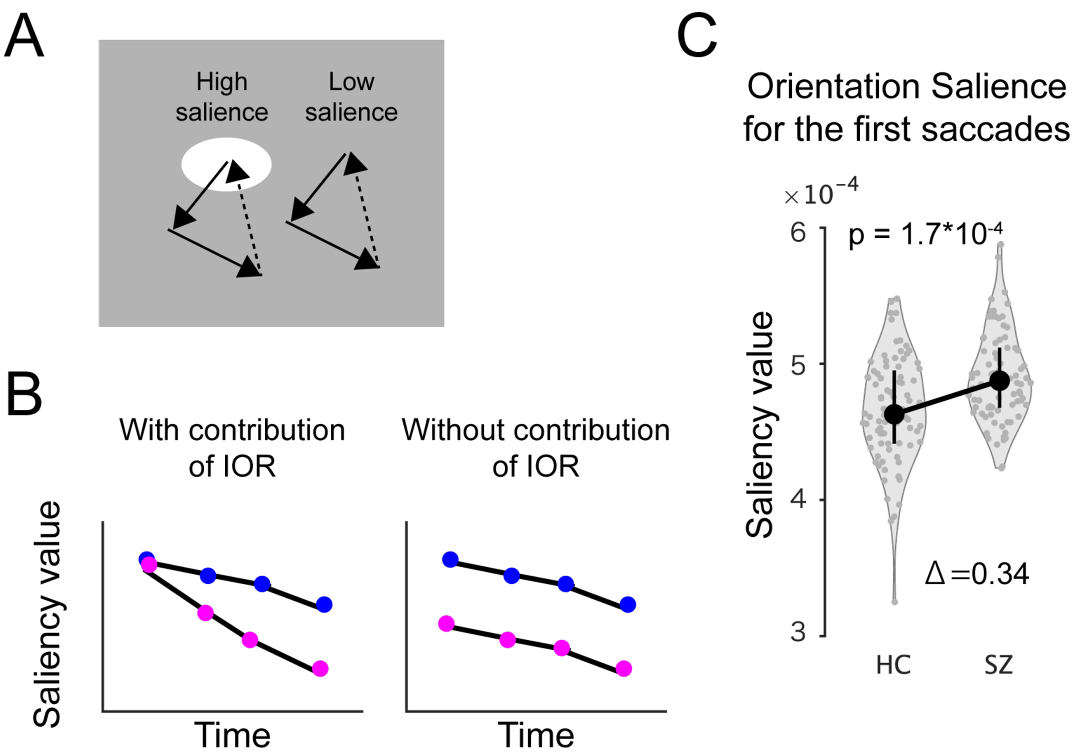


**Supplementary Figure 1. Inhibition-of-return (IOR) cannot explain the effect of orientation salience.**

(A) Possible scheme for a relationship between the return saccades (dotted line) and the visual salience of the test images (shown in brightness).

(B) Possible time courses of salience values when IOR contributes to salience-guided eye movements (left) and when IOR does not contribute to salience-guided eye movements (right). Our results in Fig. 2 support the latter.

(C) Violin plots of the mean orientation salience at the first gaze for each 8-second viewing time. The same notation as Fig. 3A.

**Supplementary Table**

| | Subjects with schizophrenia (n = 82) | Age-matched healthy controls (n = 82) | Statistics | |
|---|---|---|---|---|
| | Mean ± SD | Mean ± SD | P value | Effect size |
| Sex (male/female) | 42 / 40 | 35 / 47 | 0.28 | 0.09 |
| Age (years) | 35.1 ± 12.4 | 35.2 ± 12.3 | 0.94 | -0.01 |
| Education (years) | 13.8 ± 2.6 | 15.1 ± 2.1 | $\mathbf{5.0\times10^{-4}}$ | 0.30 |
| **Symptom-related** | | | | |
| Onset age (years) | 24.4 ± 11.4 | NA | NA | NA |
| Duration of illness (years) | 10.7 ± 8.9 | NA | NA | NA |
| CPZ equivalent (mg/day) | 600.6 ± 545.2 | NA | NA | NA |
| Cognitive decline | 13.4 ± 12.8 | NA | NA | NA |
| PANSS positive symptoms | 19.1 ± 6.1 | NA | NA | NA |
| PANSS negative symptoms | 20.8 ± 6.1 | NA | NA | NA |
| PANSS general psychopathology | 43.8 ± 12.0 | NA | NA | NA |
| PANSS total | 83.7 ± 23.2 | NA | NA | NA |
| **Cognitive tests** | | | | |
| WAIS-3 FIQ | 88.0 ± 17.9 | 112.6 ± 10.2 | $\mathbf{8.1\times10^{-18}}$ | 0.81 |
| WAIS-3 VC | 94.3 ± 17.2 | 112.5 ± 12.9 | $\mathbf{8.5\times10^{-12}}$ | 0.71 |
| WAIS-3 PO | 88.1 ± 18.0 | 106.1 ± 11.6 | $\mathbf{1.2\times10^{-10}}$ | 0.67 |
| WAIS-3 WM | 89.6 ± 17.2 | 111.1 ± 14.1 | $\mathbf{1.4\times10^{-13}}$ | 0.76 |
| WAIS-3 PS | 82.3 ± 18.3 | 110.1 ± 12.4 | $\mathbf{3.7\times10^{-18}}$ | 0.87 |
| WMS-R VerM | 89.0 ± 21.5 | 114.8 ± 13.6 | $\mathbf{1.7\times10^{-13}}$ | 0.73 |
| WMS-R ViM | 86.3 ± 18.9 | 105.5 ± 8.2 | $\mathbf{1.7\times10^{-12}}$ | 0.70 |
| WMS-R GM | 86.4 ± 21.6 | 114.0 ± 12.4 | $\mathbf{2.0\times10^{-15}}$ | 0.78 |
| WMS-R AC | 96.3 ± 15.0 | 112.0 ± 10.9 | $\mathbf{1.5\times10^{-10}}$ | 0.64 |
| WMS-R DR | 83.7 ± 21.7 | 109.7 ± 12.2 | $\mathbf{8.0\times10^{-14}}$ | 0.74 |
| UPSA-B social | 68.7 ± 16.9 | 83.0 ± 10.0 | $\mathbf{1.2\times10^{-08}}$ | 0.55 |
| SFS | 101.2 ± 31.0 | 148.5 ± 19.6 | $\mathbf{3.5\times10^{-19}}$ | 0.85 |
| Work hours | 13.0 ± 17.2 | 36.2 ± 16.9 | $\mathbf{1.4\times10^{-13}}$ | 0.71 |
| **Saccade related properties** | | | | |
| FV MainSeq a | 481.1 ± 116.5 | 452.6 ± 101.1 | 0.09 | -0.15 |
| FV MainSeq b | 9.8 ± 4.9 | 9.7 ± 3.1 | 0.55 | 0.05 |
| FV MainSeq c | 28.7 ± 9.8 | 28.4 ± 7.6 | 0.84 | 0.02 |
| FV Sac number | 16.1 ± 5.0 | 21.3 ± 4.6 | $\mathbf{9.5\times10^{-11}}$ | 0.59 |
| FV Sac amplitude | 2.9 ± 1.2 | 3.3 ± 0.8 | **0.02** | 0.21 |
| FV Scanpath Length | 72.6 ± 29.1 | 109.5 ± 22.9 | $\mathbf{5.9\times10^{-14}}$ | 0.68 |

**Supplementary table 1. Demographic data**

The “Statistics” column denotes statistical tests for the difference between SZs and healthy controls. All P values are the results of the Wilcoxon rank-sum test for two independent samples, except for the “sex” row, where a $\chi^2$ test for a 2 x 2 contingency table is performed. P values in bold denote significant differences after correction of multiple comparisons by FDR ($P < 0.05$). Cliff’s delta is shown as the effect size. Cramer’s V is shown as the effect size for the $\chi^2$ test. PANSS: Positive and negative syndrome scale. WAIS-3: Wechsler Adult Intelligence Scale 3rd edition; WMS-R: Wechsler Memory Scale-Revised; UPSA-B: UCSD Performance-based Skills Assessment - Brief; SFS: Social Functioning Scale; FV: free-viewing; Sac: saccade. MainSeq a, b, and c: parameters obtained from fitting to the main sequence relationship. See Methods for details.
